# Association between Preventive Measures against Workplace Infection and Preventive Behavior against Personal Infection

**DOI:** 10.1101/2021.06.08.21258584

**Authors:** Mika Kawasumi, Tomohisa Nagata, Hajime Ando, Ayako Hino, Seiichiro Tateishi, Mayumi Tsuji, Shinya Matsuda, Yoshihisa Fujino, Koji Mori, for the CORoNaWork project

## Abstract

**Objectives:** To prevent the spread of coronavirus disease 2019 (COVID-19) infection, it is necessary for each individual to adopt infection prevention behavior. We investigated the effect of infection control measures implemented in the workplace on personal infection prevention behavior.

**Methods:** We conducted a self-administered questionnaire survey through the Internet from December 22 to 25, 2020, during which period COVID-19 was spreading. Among respondents aged 20 to 65 years (n=27,036), a total of 21,915 workers were included in the analysis after excluding self-employed workers (n=2,202), workers in small/home offices (n=377), and agriculture, forestry, and fisheries workers (n=212), etc., whose personal infection prevention behavior was almost the same as infection control measures taken in the workplace.

**Results:** The results showed that as the number of infection control measures in the workplace increased, implementation of infection prevention behavior by individuals also significantly increased. However, the relationship differed depending on the type of personal infection prevention behavior. Specifically, infection control measures against COVID-19 in the workplace may affect personal infection prevention behavior.

**Conclusion:** Implementation of infection control measures in the workplace appears to deepen personal understanding of infection prevention behaviors, and increases awareness of the importance of individual infection prevention behavior and its implementation by all individuals. These findings may be applicable not only to COVID-19 measures but also to responses to other emerging infections and seasonal influenza.

## Introduction

An outbreak of coronavirus disease 2019 (COVID-19) occurred in Wuhan, China in December 2019, and the World Health Organization (WHO) declared a “Public Health Emergency of International Concern” on January 30, 2020.^1^ The WHO recommends taking all measures, such as avoiding the 3Cs (closed spaces, crowded places, and close-contact settings), wearing a mask, and opening windows, to protect the individual and others from COVID-19. In order to prevent the spread of the infection, it is necessary not only to take measures nationally and regionally, such as prohibiting travel or outings around the world, but also to ensure that infection prevention behavior is implemented at an individual level.^2^

Workplaces where many workers share the same space are generally vulnerable to the spread of infectious diseases. Therefore, from the perspective of business continuity, companies need to actively adopt telework, in accordance with the guidelines issued by the government and related organizations and checklists of the specific infection control measures, etc. If telework is not possible, it is necessary to implement feasible infection control measures to prevent infection in the workplace, in addition to basic measures (social distancing, wearing a mask, washing hands).^3^ However, disease clusters in the workplace are still being reported: for an approximately one-month period around December 2020, the Japanese media reported a total of 95 workplace-related disease clusters with five or more people, involving 1,103 persons in Japan.^4^ Reasons for these disease clusters were eating while talking face-to-face without wearing a mask, staying in a poorly ventilated room, seating close to other workers, and failure to disinfect shared items and equipment, etc.^5^

However, even if the infection control measures in the workplace are properly implemented, infection cannot be completely prevented if workers do not adopt appropriate infection prevention behavior, or if infection prevention behavior outside the workplace is not appropriate. It has been demonstrated that the establishment of a safe and healthy work environment under a policy of health maintenance and promotion in the workplace has a positive impact on the health of individual workers,^6-8^ and this may also be applicable to infection control measures for COVID-19. In other words, good infection control measures in the workplace may lead to personal prevention behavior against infection among workers. It has been reported that training managers is an effective and efficient way to educate workers about health issues in the workplace.^9^ Furthermore, it has been reported that when managers reach out to their workers, influenza vaccination rates increase.^10^

In a survey of Chinese occupational fields, China at the time of a factory restart after lockdown for COVID-19, it was reported that the number of preventive measures implemented by the factory in Shenzhen was associated with self-reported compliance with all four personal preventive measures (wearing a face mask consistently in any public space; sanitizing hands every time after returning from public spaces or touching installations; avoiding social and meal gatherings with people who do not live together; and avoiding crowded places).^11^ Since this survey only included factory workers in one city in China at the time of factory restart after a lockdown in which strict measures were imposed, further investigations were deemed necessary to generalize the association between workplace infection control measures and workers’ personal infection prevention behavior. Here therefore, we conducted an Internet survey during the period when COVID-19 infection was spreading in Japan and examined the relationship between infection control measures in the workplace and personal infection prevention behavior.

## Materials and Methods

A research group from the University of Occupational and Environmental Health, Japan, conducted a prospective cohort study, known as the Collaborative Online Research on Novel-coronavirus and Work study (CORoNaWork study), as a self-administered questionnaire survey via an internet survey company (Cross Marketing Inc.; Tokyo, Japan). During the baseline survey, conducted from December 22 to 25, 2020, Japan was in the midst of its third wave of the pandemic, at which point the number of COVID-19 infections and deaths was markedly higher than in the first and second waves, and the country was accordingly on high alert.

A portion of the baseline survey responses from the CORoNaWork study was used to conduct the present cross-sectional study. The study protocol, including the sampling plan and subject recruitment procedure, has been reported elsewhere in detail. Participants were aged 20-65 years who were working at the time of the baseline survey (n = 33,087 total). For participation in the CORoNaWork study, they were stratified by disease cluster sampling for gender, age, region, and occupation. After excluding 6,051 initial subjects who provided invalid responses, we included 27,036 in the database.

The flow diagram of this study is shown in Figure 1. Respondents were asked to answer yes/no to the following two questions about COVID-19 infection: “Have you had COVID-19?” and “Have you come in close contact with a person infected with COVID-19?” Those who answered yes to either question were excluded from the study (n=399). We also excluded self-employed workers (2,202), workers in small/home offices (377) and those in agriculture, forestry, and fishing (212) because of the need to evaluate infection control measures in the workplace. Workplaces with four or fewer employees were excluded from the analysis because it can be assumed that infection control measures in the workplace and personal infection prevention behavior were closely similar (one employee (n=2524) and 2-4 employees (n=1974)). We finally analyzed 21,915 workers.

### Assessment of infection control measures in the workplace

For each of the following nine infection control measures in the workplace, respondents were asked to choose whether or not the measures were being implemented: (1) prohibition/restriction of business trips; (2) prohibition/restriction of visitors; (3) prohibition of holding or limiting the number of people participating in social gatherings and banquets; (4) restriction on face-to-face meetings; (5) requirement to always wear masks during working hours; (6) installation of partitions and change of workplace layout; (7) recommendation for daily temperature check; (8) recommendation to telecommute; and (9) Request not to come to work when sick. Implementation was classified into the five categories of 0, 1-2, 3-4, 5-7, and 8-9 measures implemented.

### Assessment of personal infection prevention behavior

For each of the following seven personal infection prevention behaviors, respondents were asked to select from among four options (almost always; almost often; not often; or almost never) of how often they had performed the behavior in the last month: (1) wearing a mask in the presence of others; (2) disinfecting hands with alcohol before going indoors; (3) washing hands after using the toilet; (4) gargling when returning home; (5) opening windows and doors to ventilate the room; (6) carrying alcohol disinfectant when going out; and (7) disinfecting hands and washing hands after touching things that many people have touched. We created a binary variable by defining almost always as having good behavior, and the other responses as not having good behavior.

### Assessment of covariates

Covariates included demographics, socioeconomic factors, occupation and number of employees in the workplace. Age was expressed as a continuous variable. Yearly household income was classified into four categories: <2.50 million Japanese yen (JPY); ≥2.50 and <3.75 million JPY; ≥3.75 and <5.00 million JPY; and ≥5.00 million JPY. Education was classified into four categories: junior high school or high school, vocational school, junior college or technical school, and university or graduate school. Marital status was classified into three categories: married; divorce or widowed; or unmarried. In this survey, participants chose 1 occupation from among 10 options: general employee; manager; executive manager; public employee, faculty member, or non-profit organization employee; temporary or contract employee; self-employed; small office home office (SOHO); agriculture, forestry, or fishing; professional occupation (lawyer, tax accountant, medical-related, etc.); and other occupations. Three of these categories were excluded from this study, as mentioned above, so occupation was ultimately classified into seven categories. The number of employees in the workplace was classified into four categories: 5-9, 10-99, 100-999, and ≥1000. In addition, the cumulative incidence rate of COVID-19 infection one month prior to the conduct of the survey in the prefecture of residence was used as a community-level variable. Information was collected from the websites of public institutions.

### Statistical analyses

The odds ratios (ORs) of having good personal infection prevention behavior associated with infection control measures in the workplace were estimated using a multilevel logistic model nested in the prefectures of residence. An analysis was conducted for each of the seven personal infection prevention behaviors. The multivariate model was adjusted for age and sex, income (by category), educational background (by category), marital status, occupation and number of employees in the workplace (by category). The incidence rate of COVID-19 by prefecture was also used as a prefecture-level variable. A trend test was performed by conducting the same analysis for each of the seven personal infection prevention behaviors in the workplace using the number of infection control measures in the workplace (0-9) as a continuous variable. A p-value less than 0.05 was considered statistically significant. All analyses were conducted using Stata (Stata Statistical Software release 16; StataCorp LLC, TX, USA).

The present study was approved by the Ethics Committee of the University of Occupational and Environmental Health, Japan (reference No. R2-079 and R3-006). Informed consent was obtained from all participants.

## Results

Table 1 shows participant characteristics by category of the number of infection control measures in the workplace. Of the 21,915 participants, 7,484 (34%) were in a workplace with eight or nine infection control measures, and 1,313 (6%) were in a workplace with no infection control measures.

**Table 1.** Participants’ characteristics by category of infection control measures in the workplace

|  | Number of infection control measures in the workplace |  |  |  |  |
| --- | --- | --- | --- | --- | --- |
|  | 0 | 1-2 | 3-4 | 5-7 | 8-9 |
| Number of subjects | 1313 | 1984 | 3263 | 7871 | 7484 |
| Age, mean (SD) | 46.7 (10.2) | 46.1 (10.1) | 46.0 (10.5) | 46.2 (10.7) | 46.5 (10.6) |
| Sex, Men | 736 (56.1%) | 988 (49.8%) | 1558 (47.7%) | 3757 (47.7%) | 3861 (51.6%) |
| Equivalent income (million JPY) |  |  |  |  |  |
| <2.50 | 411 (31.3%) | 522 (26.3%) | 743 (22.8%) | 1333 (16.9%) | 970 (13.0%) |
| ≥2.50 and <3.75 | 392 (29.9%) | 663 (33.4%) | 1056 (32.4%) | 2265 (28.8%) | 1861 (24.9%) |
| ≥3.75 and <4.99 | 289 (22.0%) | 444 (22.4%) | 747 (22.9%) | 2103 (26.7%) | 2005 (26.8%) |
| ≥4.99 | 221 (16.8%) | 355 (17.9%) | 717 (22.0%) | 2170 (27.6%) | 2648 (35.4%) |
| Educational background |  |  |  |  |  |
| Junior high or high school | 556 (42.3%) | 737 (37.1%) | 1041 (31.9%) | 1999 (25.4%) | 1528 (20.4%) |
| Vocational school, junior college or technical school | 303 (23.1%) | 508 (25.6%) | 832 (25.5%) | 1899 (24.1%) | 1579 (21.1%) |
| University | 418 (31.8%) | 697 (35.1%) | 1263 (38.7%) | 3570 (45.4%) | 3758 (50.2%) |
| Graduate School | 36 (2.7%) | 42 (2.1%) | 127 (3.9%) | 403 (5.1%) | 619 (8.3%) |
| Marital status |  |  |  |  |  |
| married | 652 (49.7%) | 1012 (51.0%) | 1671 (51.2%) | 4439 (56.4%) | 4617 (61.7%) |
| widowed/divorced | 149 (11.3%) | 249 (12.6%) | 367 (11.2%) | 833 (10.6%) | 630 (8.4%) |
| unmarried | 512 (39.0%) | 723 (36.4%) | 1225 (37.5%) | 2599 (33.0%) | 2237 (29.9%) |
| Occupation |  |  |  |  |  |
| General employee | 947 (72.1%) | 1311 (66.1%) | 1793 (54.9%) | 3899 (49.5%) | 3741 (50.0%) |
| Manager | 95 (7.2%) | 146 (7.4%) | 249 (7.6%) | 795 (10.1%) | 1157 (15.5%) |
| Executive manager | 27 (2.1%) | 69 (3.5%) | 104 (3.2%) | 171 (2.2%) | 179 (2.4%) |
| Public employee, faculty member, or non-profit organization employee | 56 (4.3%) | 119 (6.0%) | 418 (12.8%) | 1302 (16.5%) | 824 (11.0%) |
| Temporary/contract employee | 132 (10.1%) | 245 (12.3%) | 507 (15.5%) | 1012 (12.9%) | 906 (12.1%) |
| Professional occupation (lawyer, tax accountant, medical-related, etc.) | 43 (3.3%) | 76 (3.8%) | 167 (5.1%) | 613 (7.8%) | 625 (8.4%) |
| Other occupation | 13 (1.0%) | 18 (0.9%) | 25 (0.8%) | 79 (1.0%) | 52 (0.7%) |
| Number of employees in the workplace |  |  |  |  |  |
| 5-9 | 273 (20.8%) | 362 (18.2%) | 369 (11.3%) | 328 (4.2%) | 146 (2.0%) |
| 10-99 | 576 (43.9%) | 998 (50.3%) | 1405 (43.1%) | 2469 (31.4%) | 1316 (17.6%) |
| 100-999 | 272 (20.7%) | 415 (20.9%) | 881 (27.0%) | 2757 (35.0%) | 2700 (36.1%) |
| ≥1000 | 192 (14.6%) | 209 (10.5%) | 608 (18.6%) | 2317 (29.4%) | 3322 (44.4%) |

Table 2 shows the proportion of each good personal infection preventive behavior and the association between category of infection control measures in the workplace and good personal infection preventive behavior. The number of infection control measures in the workplace was associated with wearing a mask in the presence of others (1-2 measures: aOR=2.12, 95% confidence interval [CI]: 1.81-2.49, p<0.001; 3-4 measures: 3.22 (2.77-3.75), p<0.001; 5-7 measures: 5.21 (4.52-6.01), p<0.001; 8-9 measures: 6.75 (5.80-7.86), p<0.001, reference to no measures). A dose-response relationship was observed in this association (p for trend; p<0.001).

**Table 2.**
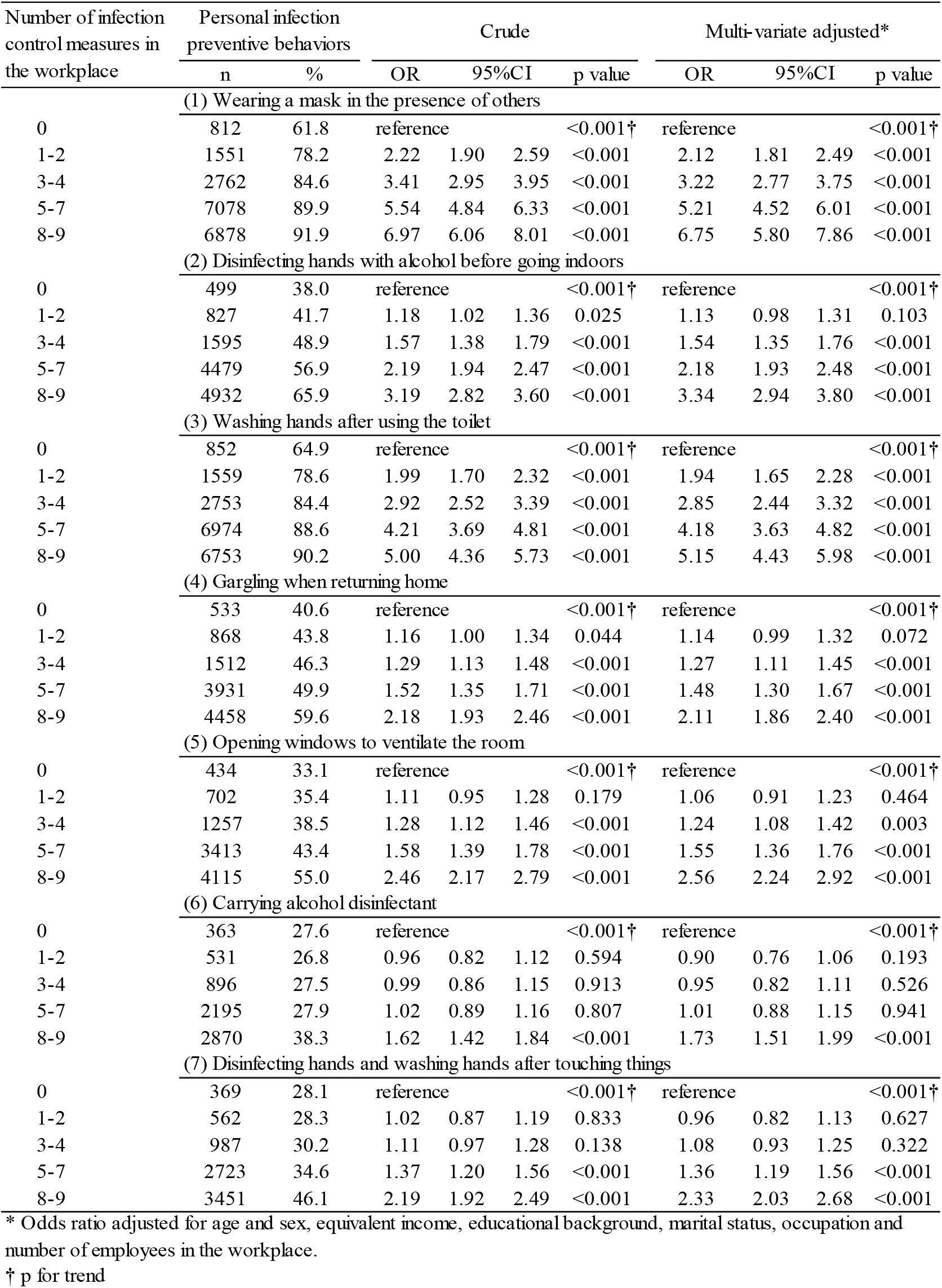
Odds ratios by number of infection control measures in the workplace and good personal infection prevention behavior

The number of infection control measures in the workplace was positively associated with carrying alcohol disinfectant when going out (1-2 measures: 0.90 (0.76-1.06), p=0.193; 3-4 measures: 0.95 (0.82-1.11), p=0.526; 5-7 measures: 1.01 (0.88-1.15), p=0.941; 8-9 measures: 1.73 (1.51-1.99), p<0.001, refer to no measures). A dose-response relationship was observed for this association (p for trend; p<0.001).

The number of infection control measures in the workplace was also associated with disinfection of hands and washing hands after touching things that many people had touched under conditions wherein infection control in the workplace is thoroughly implemented (1-2 measures: 0.96 (0.82-1.13), p=0.627; 3-4 measures: 1.08 (0.93-1.25), p=0.322; 5-7 measures: 1.36 (1.19-1.56), p<0.001; 8-9 measures: 2.33 (2.03-2.68), p<0.001, reference to no measures). A dose-response relationship was observed in this association (p for trend; p<0.001).

The number of infection control measures in the workplace was positively associated with all the good personal infection prevention behaviors (p for trend; p<0.001).

## Discussion

In this study, we classified the implementation status of infection control measures in the workplace by the number of implementation items and investigated the relationship of this classification with the implementation of personal infection prevention behavior. Results showed that implementation of infection prevention behavior by individuals significantly increased as the number of infection control measures in the workplace increased. Specifically, it was suggested that infection control measures against COVID-19 in the workplace may affect personal infection prevention behavior.

With regard to the significant relationship found between workplace infection control measures and personal infection prevention behavior, it has been reported that workplace-initiated health promotion programs encourage workers to adopt better health behavior.^6-8^ In a survey of Japanese people, it was reported that “the number of workplace measures taken in response to COVID-19 was positively associated with global fear of COVID-19.”^12^ In a survey of Ethiopian waiters, it was reported, with regard to good preventive behavior, that knowledge of COVID-19 was not high but risk perception was high.^13^ On the other hand, in a survey of Chinese occupational fields, it was reported that knowledge about transmission routes of COVID-19 was correlated with all four personal infection prevention behaviors (wearing a face mask consistently in any public space; sanitizing hands every time after returning from public spaces or touching installations; avoiding social and meal gatherings with people who do not live together; and avoiding crowded places) but the perceived severity of COVID-19 was not associated with consistent face mask wearing or sanitizing hands.^11^ Considering these previous studies, thorough implementation of infection control measures may increase knowledge and awareness of risks of COVID-19 infection, resulting in the adoption of personal infection prevention behavior. Raising awareness to change the behavior of employees is recommended as a measure of COVID-19 control in the workplace.^14,15^ Therefore, it is possible that guidance may be provided as part of infection prevention measures for individuals in those workplaces where infection control measures are actively implemented, and this may have had a direct effect on the behavior of individuals.

In this survey, the number of infection control measures in the workplace was found to be associated with good personal infection prevention behavior. The same result was also obtained in the above-mentioned survey of Chinese occupational fields.^11^ However, the association in our present survey varied according to the items of infection prevention behaviors. Compared to the situation in which the number of infection control measures in the workplace was 0, significant differences when the number of infection control measures in the workplace was 1-2 were seen wearing a mask and washing hands after using the toilet. These items were high implementation rates. Further, when there were 3-4 workplace measures, significant differences were additionally seen for alcohol hand disinfection, gargling when returning home, and room ventilation. These findings indicate that personal items that can be implemented relatively easily with a larger overall number of workplace implementations are more susceptible to the effect of the number of workplace infection control measures.

In the future, attainment of herd immunity through vaccination is considered to be the most effective way to prevent COVID-19.^16^ However, given the time required to acquire herd immunity in many countries and the impact of variants on vaccine effectiveness, it can be said that a thorough implementation of personal behavior aimed at infection prevention will continue to be an essential measure against pandemic.^17^ In Japan, even at of the end of March 2020, when the government’s request for cooperation was made, Japanese people in general had changed their behavior, indicating that understanding of the need for each individual to take action against COVID-19 was already widespread, thanks to the influence of mass media and social media.^18^ The present findings indicate that to encourage people who are not ready to change their behavior, it is important to strengthen infection control measures in the workplace where workers spend many hours of the day. In addition, it is desirable to combine multiple infection control measures, as it has been reported that the pandemic can be delayed by implementing a strategy that combines multiple measures rather than individual measures.^19^ In such cases, there are levels of difficulty in implementing infection control measures differ among different industries and job types, such as restrictions on telecommuting and interviews, and it is considered more effective to increase the number of measures to be implemented in accordance with the actual circumstances of the workplace. Although there are differences in the likelihood that the effects of workplace infection control measures influence the item of personal infection prevention behavior, it is important that workplace infection control measures are thoroughly implemented.

There are two limitations to this study. First, those who did not have Internet access or were not registered as monitors were excluded from the study. However, since subject bias was reduced by random sampling for each region/occupation/prefecture based on the incidence of infections, our results appear to be generalizable within Japan. However, since the survey was conducted in a Japanese population, generalizability to other countries appears limited. It is said that Japanese people characteristically adjust themselves to others easily,^20^ and that the implementation of infection control measures in the workplace might therefore be strongly related to personal infection prevention behavior. This study appears to support this idea, given that implementation of workplace infection control measures tended to promote personal infection prevention behavior. The findings of this study are likely to be utilized in the future not only for COVID-19 control but also for other infections, including emerging infectious diseases and seasonal influenza.

## Conclusions

We found a positive association between a greater the number of COVID-19 measures implemented in the workplace and higher likelihood of personal infection prevention behavior. This was because knowledge about effective personal infection prevention behavior is increased by implementing infection control measures in the workplace. And increased individual awareness of the risk of infection is linked to personal behavior. This in turn suggests the importance of active promotion of infection control measures in the workplace. These findings may be applicable not only to COVID-19 measures but also to responses to other emerging infections and seasonal influenza.

## Data Availability

The data that support the findings of this study are available from the corresponding author upon reasonable request.

## Acknowledgements

This study was supported and partly funded by the University of Occupational and Environmental Health, Japan; General Incorporated Foundation (Anshin Zaidan); The Development of Educational Materials on Mental Health Measures for Managers at Small-sized Enterprises; Health, Labour and Welfare Sciences Research Grants; Comprehensive Research for Women’s Healthcare (H30-josei-ippan-002); Research for the Establishment of an Occupational Health System in Times of Disaster (H30-roudou-ippan-007); and scholarship donations from Chugai Pharmaceutical Co., Ltd., the Collabo-Health Study Group, and Hitachi Systems, Ltd.

Current members of the CORoNaWork Project (in alphabetical order) are: Dr. Yoshihisa Fujino (present chairperson of the study group), Dr. Akira Ogami, Dr. Arisa Harada, Dr. Ayako Hino, Dr. Hajime Ando, Dr. Hisashi Eguchi, Dr. Kazunori Ikegami, Dr. Kei Tokutsu, Dr. Keiji Muramatsu, Dr. Koji Mori, Dr. Kosuke Mafune, Dr. Kyoko Kitagawa, Dr. Masako Nagata, Dr. Mayumi Tsuji, Ms. Ning Liu, Dr. Rie Tanaka, Dr. Ryutaro Matsugaki, Dr. Seiichiro Tateishi, Dr. Shinya Matsuda, Dr. Tomohiro Ishimaru, and Dr. Tomohisa Nagata. All members are affiliated with the University of Occupational and Environmental Health, Japan.

## Ethical approval

This study was approved by the Ethics Committee of the University of Occupational and Environmental Health, Japan (reference No. R2-079 and R3-006).

## Informed Consent

Informed consent was obtained in the form of the website.

## Registry and the Registration No. of the study/Trial

N/A

## Animal Studies

N/A

